# Key to successful treatment of COVID-19: accurate identification of severe risks and early intervention of disease progression

**DOI:** 10.1101/2020.04.06.20054890

**Authors:** Meizhu Chen, Changli Tu, Cuiyan Tan, Xiaobin Zheng, Xiaohua Wang, Jian Wu, Yiying Huang, Zhenguo Wang, Yan Yan, Zhonghe Li, Hong Shan, Jing Liu, Jin Huang

## Abstract

**Background:** COVID-19 is a new and highly contagious respiratory disease that has caused global spread, high case fatality rate in severe patients, and a huge medical burden due to invasive mechanical ventilation. The current diagnosis and treatment guidelines are still need to be improved, and more excellent clinical experience is needed to provide reference.

**Methods:** We analyzed and summarized clinical data of 97 confirmed COVID-19 adult patients (including 26 severe cases) admitted to the Fifth Affiliated Hospital of Sun Yat-sen University from January 17, 2020 to March 10, 2020, included laboratory examination results, imaging findings, treatment effect, prognosis, etc, in order to put forward prediction index of severe COVID-19 patients, principles of early intervention and methylprednisolone usages in COVID-19 patients.

**Results:**

1. Hypoxemia, hyperlactic acid, hypoproteinemia, and hypokalemia were prevalent in COVID-19 patients. The significant low lymphocyte count, hypoproteinemia, hypokalemia, the persistent or worsen high CRP, high D-dimer, and high BNP, and the occurrence of hemoptysis and novel coronavirus (SARS-CoV-2) viremia were important indicators for early diagnosis and prediction of severe disease progression.
2. Characteristic images of lung CT had a clear change in COVID-19, Ground-glass opacity (GGO) and high-density linear combinations may indicate different pathological changes. Rapid lobular progression of GGO suggests the possibility of severe disease.
3. Basic principles of early intervention treatment of COVID-19: on the premise of no effective antiviral drugs, treatment is based on supportive and symptomatic therapy (albumin supplementation, supplement of potassium, supplement blood plasma, etc.) in order to maintain the stability of the intracellular environment and adequately reactivate body immunity to clean up SARS-CoV-2.
4. According to severity, oxygenation index, body weight, age, underlying diseases, appropriate amount methylprednisolone application on severe/critical COVID-19 patients on demand, improved blood oxygen and reduced the utilization rate of invasive mechanical ventilation, case fatality rate and medical burden significantly. The most common indications for invasive mechanical ventilation should be strictly control in critical COVID-19 patients.

**Conclusions:**

1. Accurate and timely identification of clinical features in severe risks, and early and appropriate intervention can block disease progression. 2. Appropriate dose of methylprednisolone can effectively avoid invasive mechanical ventilation and reduce case fatality rate in critical COVID-19 patients.

## Introduction

COVID-19 is a new and highly contagious respiratory disease caused by Severe acute respiratory syndrome coronavirus 2 (SARS-CoV-2). Sudden and rapid progression to Acute respiratory distress syndrome (ARDS) is the severe and fatal feature of COVID-19. The current outbreak of COVID-19 has caused global pandemic. Up to March 24, 2020, there are 812,18 confirmed patients and 3,281 deaths in 31 provinces in China^[1]^. In all world, there are 1051,635 confirmed cases and 56,985 confirmed deaths^[2]^, COVID-19 has a high case fatality rate in severe patients and a huge medical burden caused by invasive mechanical ventilation^[3,4]^, National Health Commission of the People’s Republic of China has issued the relevant diagnosis and treatment guidelines for COVID-19, which have been updated to the 7th edition^[5]^, WHO also referred to the Chinese COVID-19 guidelines for diagnosis and treatment, and proposed internationally oriented advice on diagnosis and treatment^[6]^. However, with the continuous recognition of this new disease, we believe that the current diagnosis and treatment guidelines still need to be improved, such as the characteristic clinical symptoms of COVID-19, meaningful laboratory examination indicators, the diagnosis basis of covid-19 pneumonia, the limitation of glucocorticoid usage and dosage in treatment, and the choice of mechanical ventilation timing. More excellent experience in clinical diagnosis and treatment is needed for reference.

The Fifth Affiliated Hospital of Sun Yat-sen University, as the only designated COVID-19 hospital in Zhuhai city, has admitted 97 cases of COVID-19 patients, all of whom have been successfully cured and discharged. Through the analysis and summary of the diagnosis and treatment process of these patients, we hope to provide some reference advice on the diagnosis and treatment of COVID-19, especially how to accurately identify the risk of severe disease and early block disease progression, and rationally apply glucocorticoid to reduce the rate of invasive mechanical ventilation and case fatality rate of severe patients.

## Material and method

### 1. Data sources

This study was a retrospective single-center study, which included 97 confirmed COVID-19 adult patients (more than 14 years old) admitted to the Fifth Affiliated Hospital of Sun Yat-sen University from January 17, 2020 to March 10, 2020. This retrospective observational study was approved by the Research Ethics Committee of The Fifth Affiliated Hospital of Sun Yat-sen University and the need for informed consent was waived, considering the retrospective study design. Clinical data were extracted from electronic medical records, including basic demographic data, symptoms, vital signs, clinical classification, complications and the SARS-CoV-2 RNA of nasopharyngeal swab and blood samples clearance time which was defined as the time of after two consecutive negative real-time PCR results with 1-day interval.

### 2. Diagnostic basis

COVID-19 was diagnosed and clinically classified according to the new coronavirus pneumonia diagnosis and treatment plan (trial version 7)^[5]^.

### 3. Statistical analysis

Categorical variables were described as frequency rates and percentages, and continuous variables were described using mean (±SD, standard deviation), median and interquartilerange (IQR) values. Means for continuous variables were compared using independent group t tests and one way ANVOA when the data were normally distributed and homoscedasticity; otherwise, the Mann-Whitney U test and Kruskal-Wallis H were used when the data were nonnormal distribution. Proportions for categorical variables were compared using the chi-squared test, although the Fisher exact test was used when the data were limited. All Statistical analysis was performed using SPSS software (Version 26.0, IBM). For unadjusted comparisons, a two-sided P<0.05 was considered statistically significant.

## Results

### 1. Demographic and clinical characteristics on admission of COVID-19 patients

A total of 103 patients with positive SARS-CoV-2 nasopharyngeal swab test in Zhuhai, 5 of whom were younger than 14 years old, belonged to pediatric diagnosis and treatment; the first case (death case) among adults was not treated by our medical team. A total of 97 patients treated by the diagnosis and treatment team included in the analysis of this report, of which 26 were severe, accounting for 26.8%. Among the cases of COVID-19 in Zhuhai, 69 of the 97 patients were local residents of Wuhan or had lived in Wuhan within 14 days before the onset, 8 cases were from Hubei area outside Wuhan, 14 had a history of direct contact with personnel from Hubei, and another 6 cases did not have a clear epidemiological history. The proportion of severe cases of patients directly from Wuhan was lower than that of non-severe cases (P=0.028), but the proportion of severe cases of patients who had no clear epidemiological history was higher (P=0.023). This coincides with the possibility of multiple origins of the new coronavirus. Clinical symptoms found in these COVID-19 patients were: fever, shortness of breath, cough, diarrhea, loss of appetite, hemoptysis, fatigue, nasal congestion, runny nose, chest tightness, and sputum. Hemoptysis was a distinctive symptoms of severe patients (26.9% vs 0, p <0.001 table1); shortness of breath was more common in severe patients (100% vs 9.9%, p <0.001 table1), and the incidence of fever symptoms was more in severe patients than non-severe (80.8% vs 50.7%, p = 0.008 table1). There was no any clinical symptoms in 12.4% of non-severe COVID-19 patients. The underlying diseases of these patients were: hypertension in 16 cases, diabetes mellitus in six cases, *PFS* (Progression-Free-Survival)lung cancer in two cases, cured nasopharyngeal cancer in one case, cured breast cancer in one case, cured bladder cancer in one case, old tuberculosis in two cases, mental illness in two cases, hypothyroidism in two cases, coronary heart disease in two case, hyperlipidemia in one case, and renal failure in one case. There was no statistical difference between non-severe and severe patients (P> 0.05). The basic information and clinical symptoms of patients included in this study are shown in Table 1.

**Table 1.**
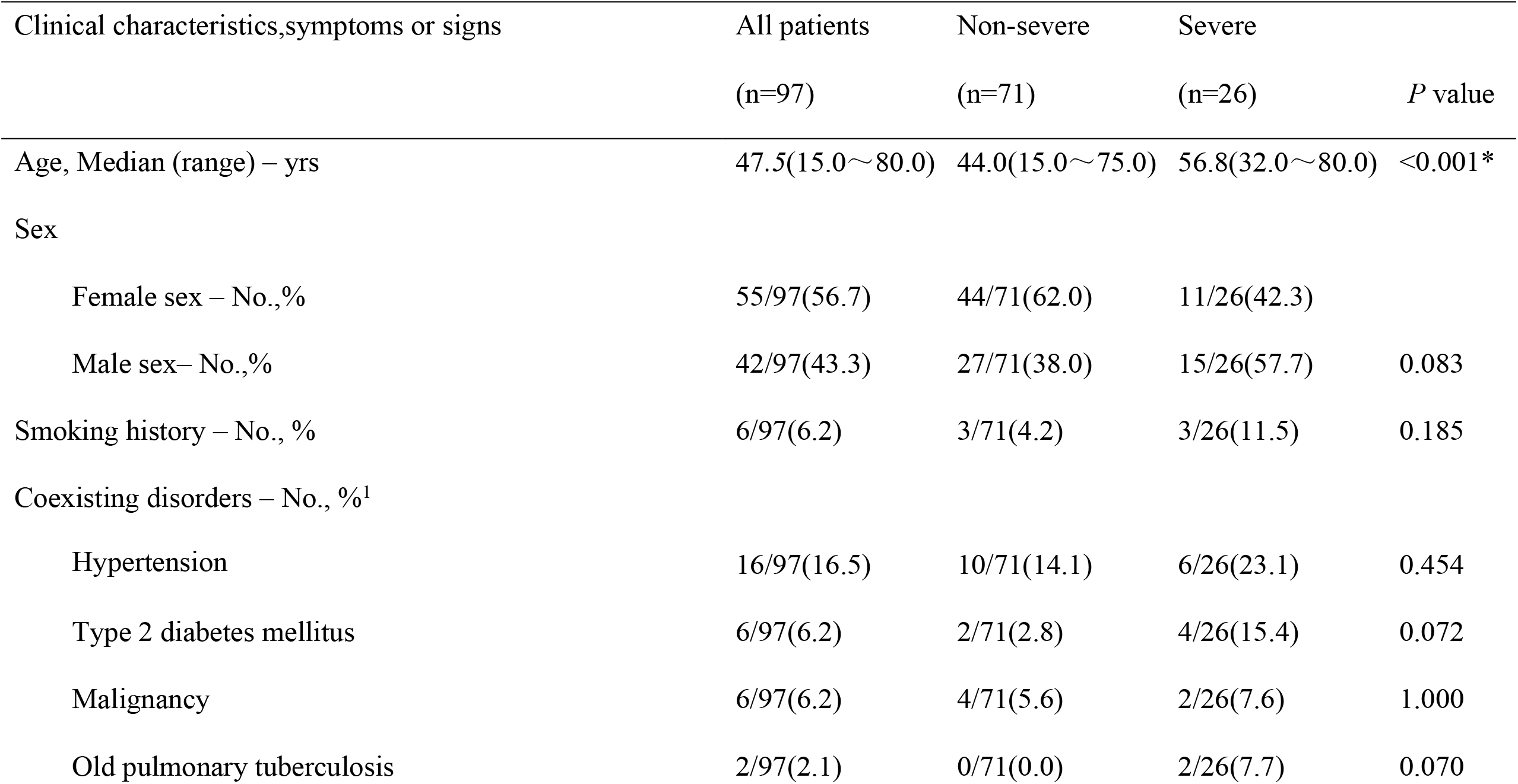
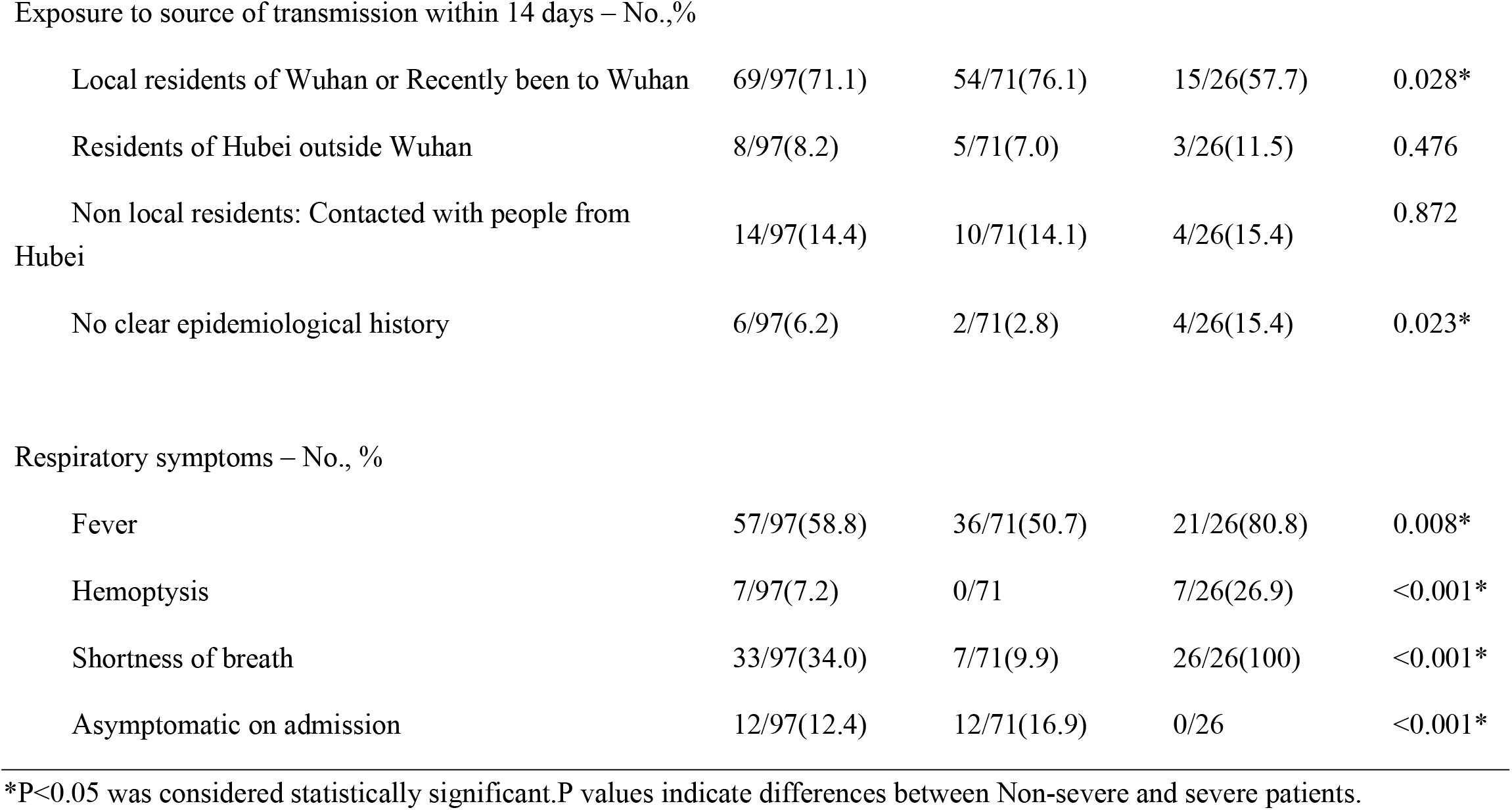
Demographic and clinical characteristics on admission of 97 patients with COVID-19

| Clinical characteristics,symptoms or signs | All patients<br>(n=97) | Non-severe<br>(n=71) | Severe<br>(n=26) | *P* value |
| --- | --- | --- | --- | --- |
| Age, Median (range) – yrs | 47.5(15.0~80.0) | 44.0(15.0~75.0) | 56.8(32.0~80.0) | <0.001* |
| Sex |  |  |  |  |
| Female sex – No.,% | 55/97(56.7) | 44/71(62.0) | 11/26(42.3) |  |
| Male sex– No.,% | 42/97(43.3) | 27/71(38.0) | 15/26(57.7) | 0.083 |
| Smoking history – No., % | 6/97(6.2) | 3/71(4.2) | 3/26(11.5) | 0.185 |
| Coexisting disorders – No., %<sup>1</sup> |  |  |  |  |
| Hypertension | 16/97(16.5) | 10/71(14.1) | 6/26(23.1) | 0.454 |
| Type 2 diabetes mellitus | 6/97(6.2) | 2/71(2.8) | 4/26(15.4) | 0.072 |
| Malignancy | 6/97(6.2) | 4/71(5.6) | 2/26(7.6) | 1.000 |
| Old pulmonary tuberculosis | 2/97(2.1) | 0/71(0.0) | 2/26(7.7) | 0.070 |
| Exposure to source of transmission within 14 days – No.,% |  |  |  |  |
| Local residents of Wuhan or Recently been to Wuhan | 69/97(71.1) | 54/71(76.1) | 15/26(57.7) | 0.028* |
| Residents of Hubei outside Wuhan | 8/97(8.2) | 5/71(7.0) | 3/26(11.5) | 0.476 |
| Non local residents: Contacted with people from Hubei | 14/97(14.4) | 10/71(14.1) | 4/26(15.4) | 0.872 |
| No clear epidemiological history | 6/97(6.2) | 2/71(2.8) | 4/26(15.4) | 0.023* |
| Respiratory symptoms – No., % |  |  |  |  |
| Fever | 57/97(58.8) | 36/71(50.7) | 21/26(80.8) | 0.008* |
| Hemoptysis | 7/97(7.2) | 0/71 | 7/26(26.9) | <0.001* |
| Shortness of breath | 33/97(34.0) | 7/71(9.9) | 26/26(100) | <0.001* |
| Asymptomatic on admission | 12/97(12.4) | 12/71(16.9) | 0/26 | <0.001* |

\*P<0.05 was considered statistically significant.P values indicate differences between Non-severe and severe patients.

### 2. Characteristics of laboratory findings during the whole course of COVID-19 patients

Routine examinations were performed in almost all 97 COVID-19 patients, including total blood count, liver and kidney function, electrolyte, blood gas analysis, LDH, CK, CK-MB,α-HBDH, CRP, BNP, D-dimer, lung CT.

We selected and analyzed the most significant abnormal changes in the relevant laboratory indicators over the course of COVID-19. Throughout the disease course, hypoxemia (93.8%), hyperlactic acidemia (89.7%), hypoproteinemia (89.7%), hypokalemia (70.1%), and high CRP(53.6%) were common in 97 COVID-19 patients. Above indicators (except lactic acid) also showed significant differences between non-severe and severe (P<0.05, Table2).

**Table2.**
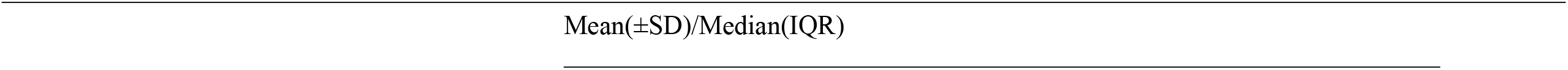
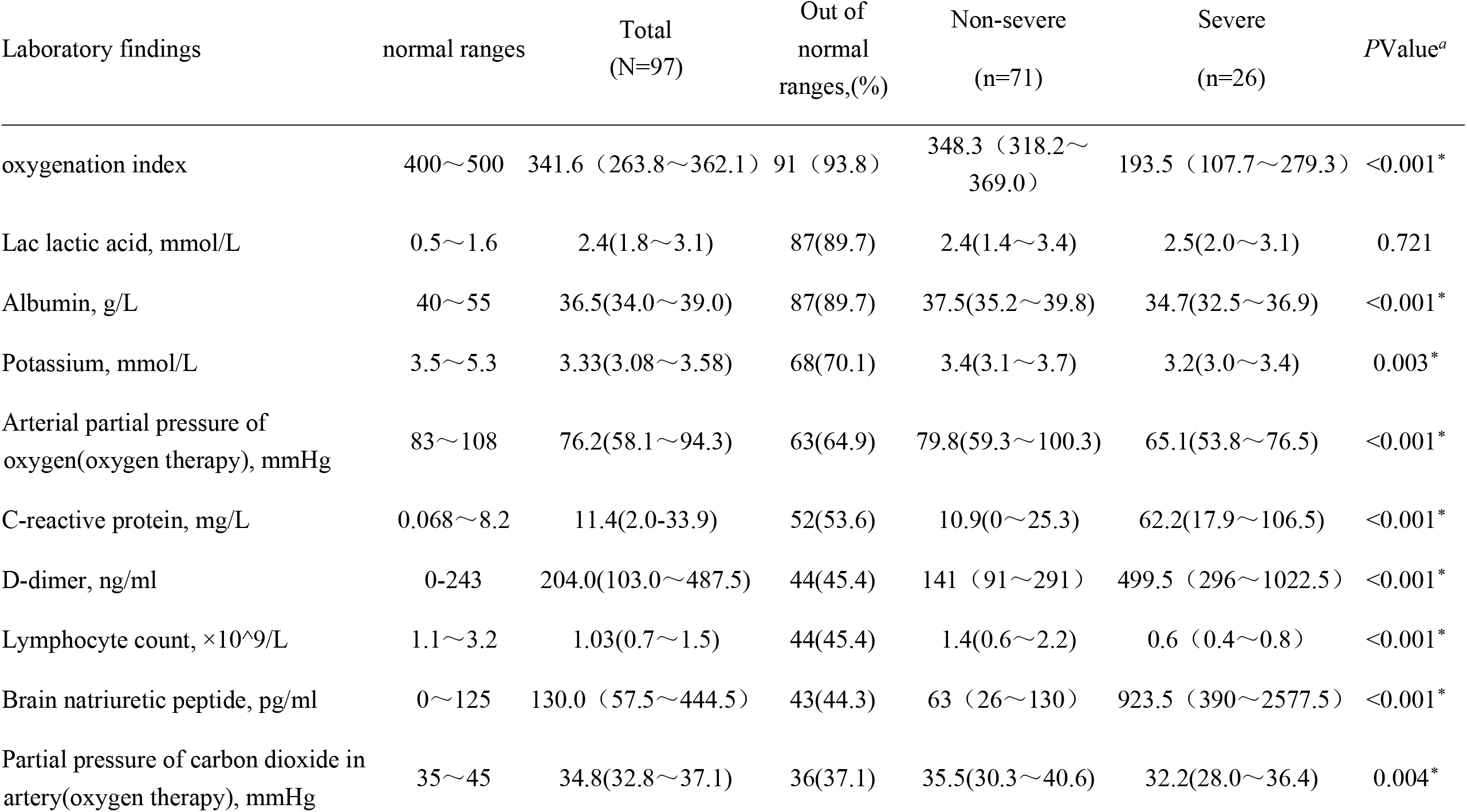
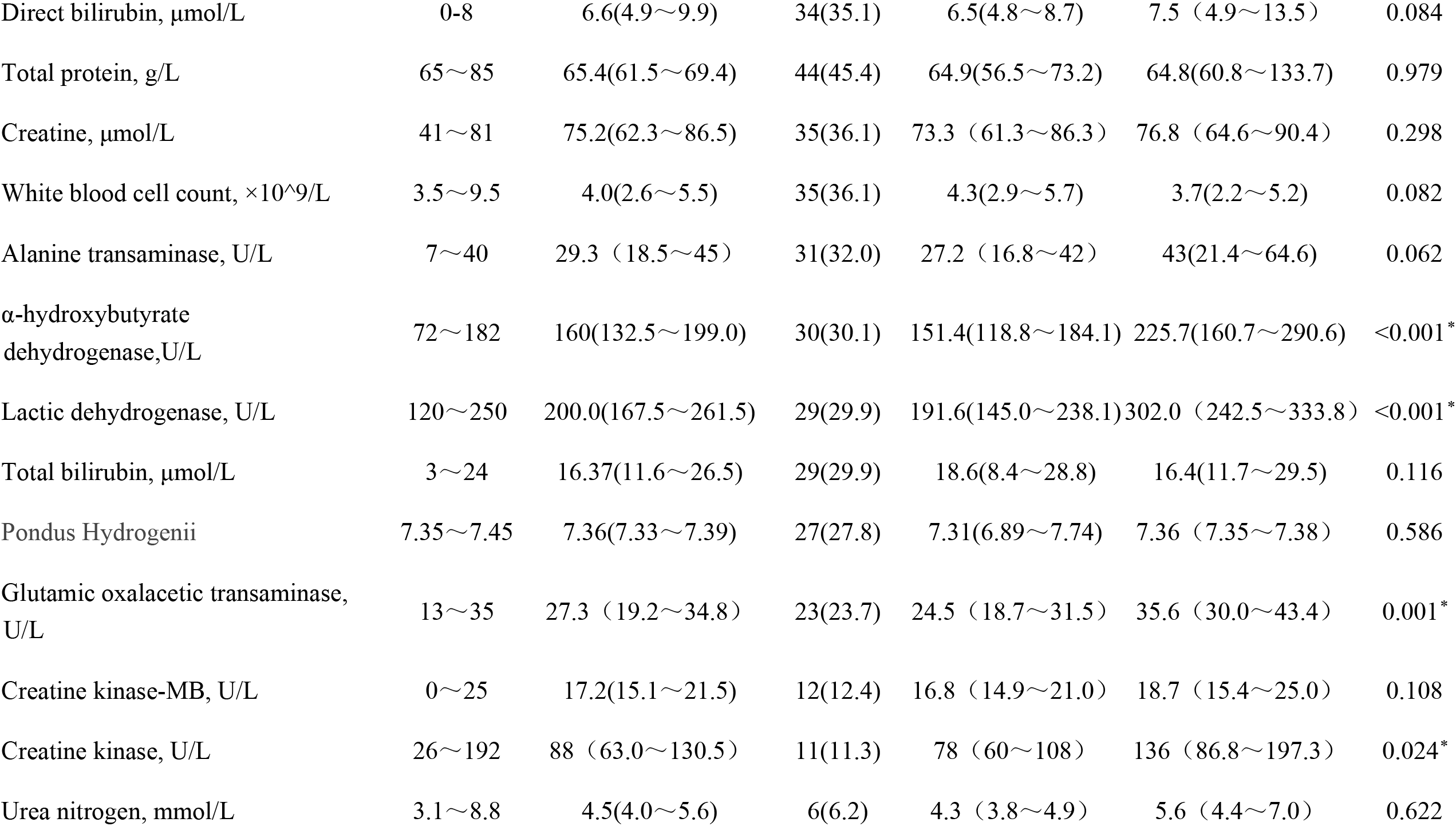
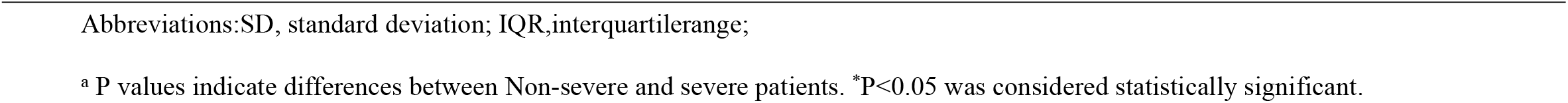
The most abnormal changes in laboratory examination in 97 COVID-19 patients

| Laboratory findings | normal ranges | Total<br>(N=97) | Out of<br>normal<br>ranges, (%) | Non-severe<br>(n=71) | Severe<br>(n=26) | PValue<sup>a</sup> |
| --- | --- | --- | --- | --- | --- | --- |
| oxygenation index | 400~500 | 341.6 (263.8~362.1) | 91 (93.8) | 348.3 (318.2~369.0) | 193.5 (107.7~279.3) | <0.001* |
| Lac lactic acid, mmol/L | 0.5~1.6 | 2.4(1.8~3.1) | 87(89.7) | 2.4(1.4~3.4) | 2.5(2.0~3.1) | 0.721 |
| Albumin, g/L | 40~55 | 36.5(34.0~39.0) | 87(89.7) | 37.5(35.2~39.8) | 34.7(32.5~36.9) | <0.001* |
| Potassium, mmol/L | 3.5~5.3 | 3.33(3.08~3.58) | 68(70.1) | 3.4(3.1~3.7) | 3.2(3.0~3.4) | 0.003* |
| Arterial partial pressure of<br>oxygen(oxygen therapy), mmHg | 83~108 | 76.2(58.1~94.3) | 63(64.9) | 79.8(59.3~100.3) | 65.1(53.8~76.5) | <0.001* |
| C-reactive protein, mg/L | 0.068~8.2 | 11.4(2.0-33.9) | 52(53.6) | 10.9(0~25.3) | 62.2(17.9~106.5) | <0.001* |
| D-dimer, ng/ml | 0-243 | 204.0(103.0~487.5) | 44(45.4) | 141 (91~291) | 499.5 (296~1022.5) | <0.001* |
| Lymphocyte count, ×10<sup>9</sup>/L | 1.1~3.2 | 1.03(0.7~1.5) | 44(45.4) | 1.4(0.6~2.2) | 0.6 (0.4~0.8) | <0.001* |
| Brain natriuretic peptide, pg/ml | 0~125 | 130.0 (57.5~444.5) | 43(44.3) | 63 (26~130) | 923.5 (390~2577.5) | <0.001* |
| Partial pressure of carbon dioxide in<br>artery(oxygen therapy), mmHg | 35~45 | 34.8(32.8~37.1) | 36(37.1) | 35.5(30.3~40.6) | 32.2(28.0~36.4) | 0.004* |
| Direct bilirubin, μmol/L | 0~8 | 6.6(4.9~9.9) | 34(35.1) | 6.5(4.8~8.7) | 7.5 (4.9~13.5) | 0.084 |
| Total protein, g/L | 65~85 | 65.4(61.5~69.4) | 44(45.4) | 64.9(56.5~73.2) | 64.8(60.8~133.7) | 0.979 |
| Creatine, μmol/L | 41~81 | 75.2(62.3~86.5) | 35(36.1) | 73.3 (61.3~86.3) | 76.8 (64.6~90.4) | 0.298 |
| White blood cell count, ×10<sup>9</sup>/L | 3.5~9.5 | 4.0(2.6~5.5) | 35(36.1) | 4.3(2.9~5.7) | 3.7(2.2~5.2) | 0.082 |
| Alanine transaminase, U/L | 7~40 | 29.3 (18.5~45) | 31(32.0) | 27.2 (16.8~42) | 43(21.4~64.6) | 0.062 |
| α-hydroxybutyrate<br>dehydrogenase,U/L | 72~182 | 160(132.5~199.0) | 30(30.1) | 151.4(118.8~184.1) | 225.7(160.7~290.6) | <0.001* |
| Lactic dehydrogenase, U/L | 120~250 | 200.0(167.5~261.5) | 29(29.9) | 191.6(145.0~238.1) | 302.0 (242.5~333.8) | <0.001* |
| Total bilirubin, μmol/L | 3~24 | 16.37(11.6~26.5) | 29(29.9) | 18.6(8.4~28.8) | 16.4(11.7~29.5) | 0.116 |
| Pondus Hydrogenii | 7.35~7.45 | 7.36(7.33~7.39) | 27(27.8) | 7.31(6.89~7.74) | 7.36 (7.35~7.38) | 0.586 |
| Glutamic oxalacetic transaminase,<br>U/L | 13~35 | 27.3 (19.2~34.8) | 23(23.7) | 24.5 (18.7~31.5) | 35.6 (30.0~43.4) | 0.001* |
| Creatine kinase-MB, U/L | 0~25 | 17.2(15.1~21.5) | 12(12.4) | 16.8 (14.9~21.0) | 18.7 (15.4~25.0) | 0.108 |
| Creatine kinase, U/L | 26~192 | 88 (63.0~130.5) | 11(11.3) | 78 (60~108) | 136 (86.8~197.3) | 0.024* |
| Urea nitrogen, mmol/L | 3.1~8.8 | 4.5(4.0~5.6) | 6(6.2) | 4.3 (3.8~4.9) | 5.6 (4.4~7.0) | 0.622 |

Abbreviations:SD, standard deviation; IQR,interquartilerange;
<sup>a</sup> P values indicate differences between Non-severe and severe patients. \*P<0.05 was considered statistically significant.

In addition, the valley value of lymphocyte count and PCO2 was lower, while the peak value of BNP and the D-dimer were higher in severe COVID-19 patients than the non-severe ones(P<0.05, Table2).

In addition, we found the proportion of abnormal of Lactic dehydrogenase, Total protein, Pondus Hydrogenii, Glutamic oxalacetic transaminase, Creatine kinase MB, Creatine kinase, Urea nitrogen was low (< 30%, Table2) in these patients. The abnormal of Glutamic oxalacetic transaminase, Lactic dehydrogenase and Creatine kinase MB tend to occur in severe COVID-19 patients (P<0.05, Table2).

### 3. Significant laboratory and clinical predictors of severe/critical COVID-19

For the above indicators with significant differences in COVID-19, we are more concerned about their changes in the course of disease, especially for the prediction of severe disease. Lymphocyte count, CRP, D-dimer, BNP, albumin, and potassium were abnormal throughout most of the course of the severe COVID-19 patients (Figure 1). It suggested that abnormalities in these indicators can early predict the progression of the severe disease. Lymphocyte and CRP recovery were slow in severe patients (Figure 1). So it must be attention to the balance between early immune response and lateral inflammatory reaction, especially during the first 2 weeks of the course of the disease. The level of D-dimer and BNP gradually increased during the first week of the course of the disease, and reached the peak at day 8-9 of the course of the disease, and then slowly decreased, which suggested that the risk of secondary thrombosis and myocardial injury should be paid attention to during the peak period. The recovery of hypoalbuminemia was still slow in the case of continuous supplementation (20g-40g albumin per day), lasting the whole course of severe patients, which also suggested that the consumption and loss of albumin may persist. Hypokalemia was also more obvious in severe patients, but it can be basically maintained in the normal range after active potassium supplementation. The overall course of the disease in severe patients lasted about three weeks before most indicators return to normal levels(Figure 1).

**Figure 1:**
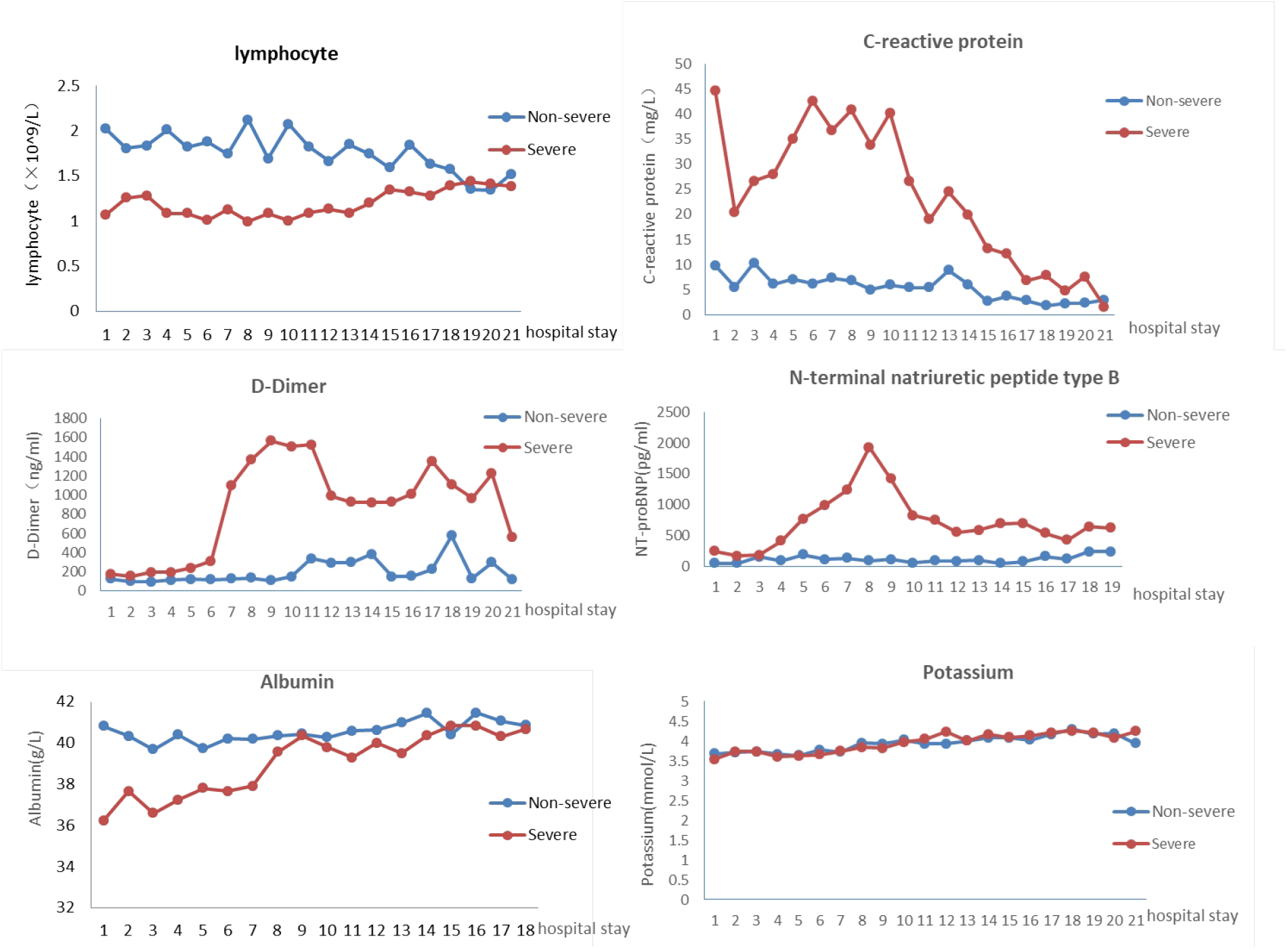
Comparsion charts of significant laboratory indicators. The curves of albumin and serum potassium were all plotted after the treatment of albumin supplementation and potassium supplementation.

As shown in Table3, COVID-19 patients were classified as mild (OI> 300mmHg), severe (150≤OI≤300mmHg), critical (OI<150mmHg) stage. Among the 26 severe patients, three patients were severe on admission, the other 23 patients all progressed rapidly from non-severe to severe stage, and 10 cases developed to critical stage with oxygenation index further decreased to less than 150mmHg.

**Table3:**
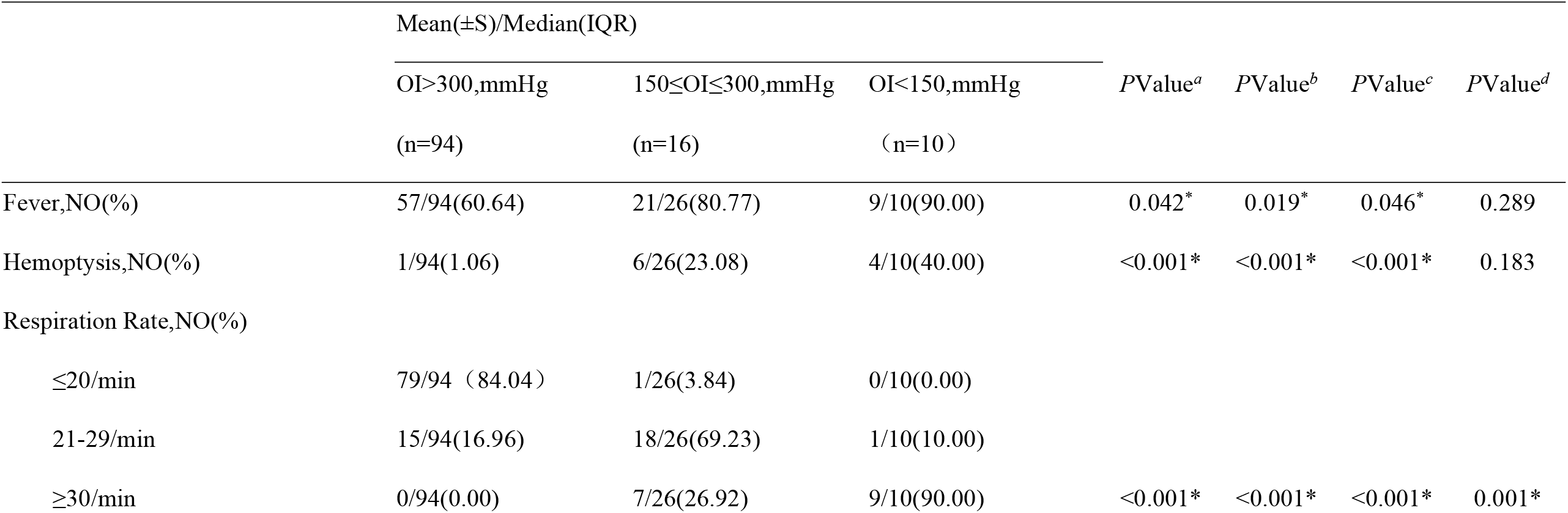
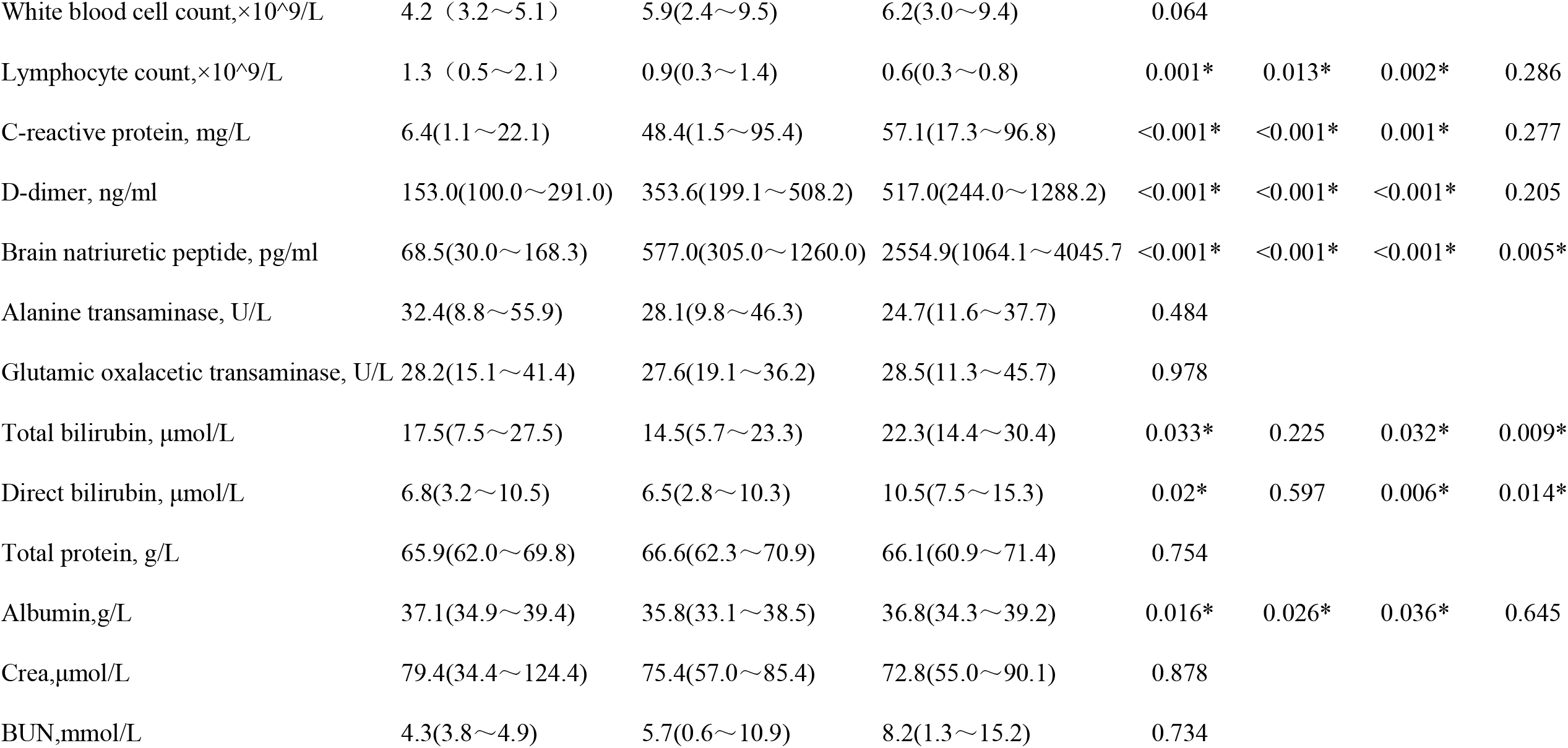
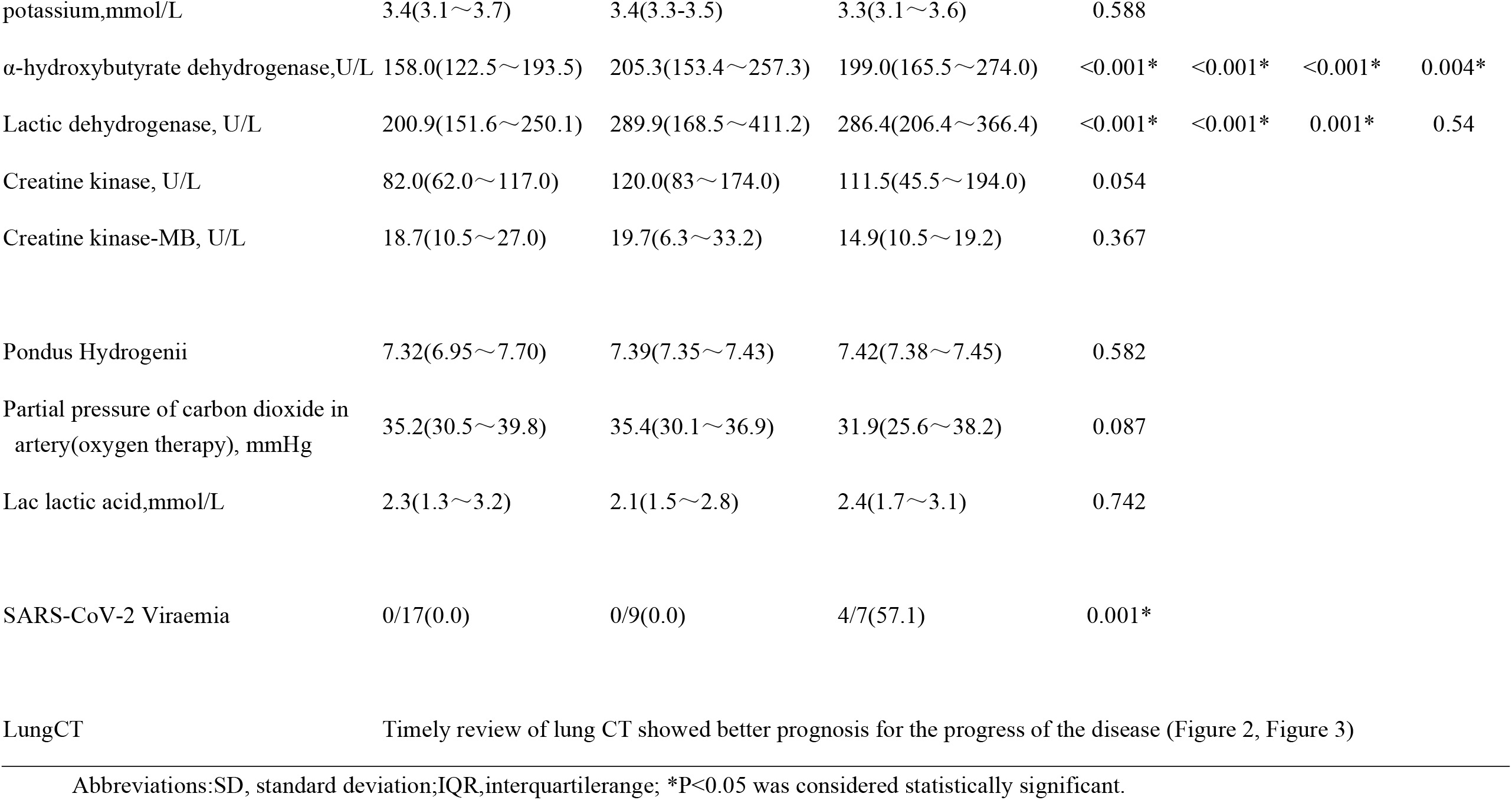
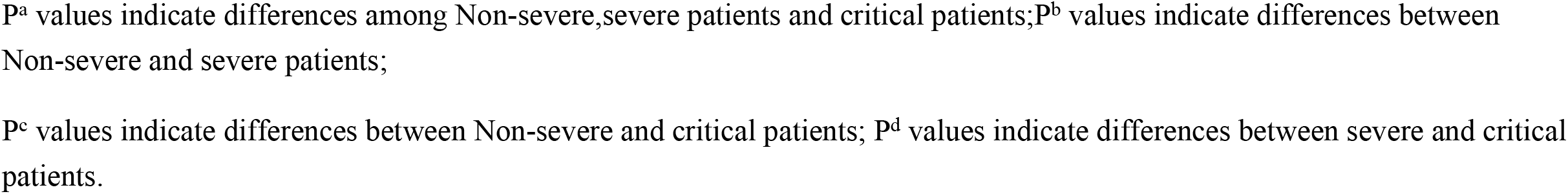
Clinical changes in COVID-19 patients

|  | Mean(±S)/Median(IQR) |  |  | PValue<sup>a</sup> | PValue<sup>b</sup> | PValue<sup>c</sup> | PValue<sup>d</sup> |
| --- | --- | --- | --- | --- | --- | --- | --- |
|  | OI>300,mmHg | 150≤OI≤300,mmHg | OI<150,mmHg |  |  |  |  |
|  | (n=94) | (n=16) | (n=10) |  |  |  |  |
| Fever,NO(%) | 57/94(60.64) | 21/26(80.77) | 9/10(90.00) | 0.042* | 0.019* | 0.046* | 0.289 |
| Hemoptysis,NO(%) | 1/94(1.06) | 6/26(23.08) | 4/10(40.00) | <0.001* | <0.001* | <0.001* | 0.183 |
| Respiration Rate,NO(%) |  |  |  |  |  |  |  |
| ≤20/min | 79/94 (84.04) | 1/26(3.84) | 0/10(0.00) |  |  |  |  |
| 21-29/min | 15/94(16.96) | 18/26(69.23) | 1/10(10.00) |  |  |  |  |
| ≥30/min | 0/94(0.00) | 7/26(26.92) | 9/10(90.00) | <0.001* | <0.001* | <0.001* | 0.001* |
| White blood cell count,×10<sup>9</sup>/L | 4.2 (3.2~5.1) | 5.9(2.4~9.5) | 6.2(3.0~9.4) | 0.064 |  |  |  |
| Lymphocyte count,×10<sup>9</sup>/L | 1.3 (0.5~2.1) | 0.9(0.3~1.4) | 0.6(0.3~0.8) | 0.001* | 0.013* | 0.002* | 0.286 |
| C-reactive protein, mg/L | 6.4(1.1~22.1) | 48.4(1.5~95.4) | 57.1(17.3~96.8) | <0.001* | <0.001* | 0.001* | 0.277 |
| D-dimer, ng/ml | 153.0(100.0~291.0) | 353.6(199.1~508.2) | 517.0(244.0~1288.2) | <0.001* | <0.001* | <0.001* | 0.205 |
| Brain natriuretic peptide, pg/ml | 68.5(30.0~168.3) | 577.0(305.0~1260.0) | 2554.9(1064.1~4045.7) | <0.001* | <0.001* | <0.001* | 0.005* |
| Alanine transaminase, U/L | 32.4(8.8~55.9) | 28.1(9.8~46.3) | 24.7(11.6~37.7) | 0.484 |  |  |  |
| Glutamic oxalacetic transaminase, U/L | 28.2(15.1~41.4) | 27.6(19.1~36.2) | 28.5(11.3~45.7) | 0.978 |  |  |  |
| Total bilirubin, μmol/L | 17.5(7.5~27.5) | 14.5(5.7~23.3) | 22.3(14.4~30.4) | 0.033* | 0.225 | 0.032* | 0.009* |
| Direct bilirubin, μmol/L | 6.8(3.2~10.5) | 6.5(2.8~10.3) | 10.5(7.5~15.3) | 0.02* | 0.597 | 0.006* | 0.014* |
| Total protein, g/L | 65.9(62.0~69.8) | 66.6(62.3~70.9) | 66.1(60.9~71.4) | 0.754 |  |  |  |
| Albumin,g/L | 37.1(34.9~39.4) | 35.8(33.1~38.5) | 36.8(34.3~39.2) | 0.016* | 0.026* | 0.036* | 0.645 |
| Crea,μmol/L | 79.4(34.4~124.4) | 75.4(57.0~85.4) | 72.8(55.0~90.1) | 0.878 |  |  |  |
| BUN,mmol/L | 4.3(3.8~4.9) | 5.7(0.6~10.9) | 8.2(1.3~15.2) | 0.734 |  |  |  |
| potassium,mmol/L | 3.4(3.1~3.7) | 3.4(3.3-3.5) | 3.3(3.1~3.6) | 0.588 |  |  |  |
| α-hydroxybutyrate dehydrogenase,U/L | 158.0(122.5~193.5) | 205.3(153.4~257.3) | 199.0(165.5~274.0) | <0.001* | <0.001* | <0.001* | 0.004* |
| Lactic dehydrogenase, U/L | 200.9(151.6~250.1) | 289.9(168.5~411.2) | 286.4(206.4~366.4) | <0.001* | <0.001* | 0.001* | 0.54 |
| Creatine kinase, U/L | 82.0(62.0~117.0) | 120.0(83~174.0) | 111.5(45.5~194.0) | 0.054 |  |  |  |
| Creatine kinase-MB, U/L | 18.7(10.5~27.0) | 19.7(6.3~33.2) | 14.9(10.5~19.2) | 0.367 |  |  |  |
| Pondus Hydrogenii | 7.32(6.95~7.70) | 7.39(7.35~7.43) | 7.42(7.38~7.45) | 0.582 |  |  |  |
| Partial pressure of carbon dioxide in artery(oxygen therapy), mmHg | 35.2(30.5~39.8) | 35.4(30.1~36.9) | 31.9(25.6~38.2) | 0.087 |  |  |  |
| Lac lactic acid,mmol/L | 2.3(1.3~3.2) | 2.1(1.5~2.8) | 2.4(1.7~3.1) | 0.742 |  |  |  |
| SARS-CoV-2 Viraemia | 0/17(0.0) | 0/9(0.0) | 4/7(57.1) | 0.001* |  |  |  |
| LungCT | Timely review of lung CT showed better prognosis for the progress of the disease (Figure 2, Figure 3) |  |  |  |  |  |  |

Abbreviations:SD, standard deviation;IQR,interquartilerange; \*P<0.05 was considered statistically significant.
P<sup>a</sup> values indicate differences among Non-severe,severe patients and critical patients;P<sup>b</sup> values indicate differences between Non-severe and severe patients;
P<sup>c</sup> values indicate differences between Non-severe and critical patients; P<sup>d</sup> values indicate differences between severe and critical patients.

We compared the characteristic symptoms and laboratory results of these groups at different stages. In addition to the oxygenation index which was an important indicator, we are more concerned about the predictive effect of changes in clinical symptoms and relevant laboratory indicators.

We also selected and analyzed the most significant abnormal changes in the relevant symptoms and laboratory indicators in three stages over the course of COVID-19. Hemoptysis, dyspnea, hyperbilirubinemia, high α-HBDH and high LDH were more common in severe and critical stage than in mild stage (P<0.05). Lymphocyte count, CRP, D-dimer, BNP, and Albumin were generally abnormal in the course of COVID-19 patients, and the abnormal changes were more significant with the aggravation of the disease. (P<0.05). The deterioration of the above indicators indicated that the patient’s condition was rapid progress, which required close monitoring and early intervention.

Blood SARS-CoV-2 RNA was detected in 33 of the 97 COVID-19 patients enrolled in this study. The results of Viral nucleic acid were interpreted as positive results in four patients, and the oxygenation index of all the 4 patients was lower than 150mmHg. It is suggested that patients with positive blood SARS-CoV nucleic acid test had a high potential risk, and are likely to progress to severe or critical disease, so the changes of the condition should be closely observed.

### 4. Characteristics of lung CT of COVID-19 and prediction of severe risk

The pneumonia like change is important lung sign in COVID-19. If there is no change in lung CT, it will not progress to severe disease. The common sign in lung CT is ground-glass opacity (GGO) and some high-density line-like and flake-like consolidation. If GGO enlarged fast or new GGO lesions appeared in multiple lobes (as shown in Figures 2-A-B), high-density line-like consolidation appeared in the central part of GGO lesions or fusioned quickly into a sheet (as shown in Figures 3-A-B), we should pay attention to the acute deterioration of COVID-19. As for lesion range on lung CT scans, we saw a greater frequency of bilateral lung involvement, and a greater number of pulmonary lobes involved, higher frequency of consolidation and more mediastinal lymph node enlargement in severe COVID-19 cases than in non-severe ones (5 vs 1, P <0.001, Table 4), suggesting that both lungs involved, multiple lobes involvement and rapid lesion progression are important indicators for predicting disease progression to severe disease.

**Table4.** Lung CT features in COVID-19 patients 3days before and after of the lowest oxygenation index

| CT imaging features | All patients<br>(n=97) | Non-severe<br>(n=71) | Severe<br>(n=26) | *P* value |
| --- | --- | --- | --- | --- |
| one or both lungs<sup>a</sup> | 2(0,2) | 1(0,2) | 2(1,2) | 0.001* |
| number of lung lobes(0-5)<sup>a</sup> | 2(0,5) | 1(0,5) | 3(1,5) | <0.001* |
| ground glass opacity ( GGO),NO(%) | 68(67.7) | 46(64.8) | 22(84.6) | 0.067 |
| Stripes or consolidation,NO(%) | 17(17.5) | 10(14.1) | 15(57.7) | <0.001* |
| nodules,NO(%) | 20(20.6) | 15(21.1) | 5(19.2) | 0.838 |
| bronchial abnormalities,NO(%) | 7(7.2%) | 3(4.2) | 4(15.4) | 0.150 |
| pleural effusion,NO(%) | 1(1.5%) | 0(0) | 1(3.8) | 0.268 |
| Others (emphysema/old tuberculosis, etc),NO(%) | 27(27.8) | 16(22.5) | 11(42.3) | 0.054 |

\*P<0.05 was considered statistically significant. a was median(min,max) values.

**Figure 2:**
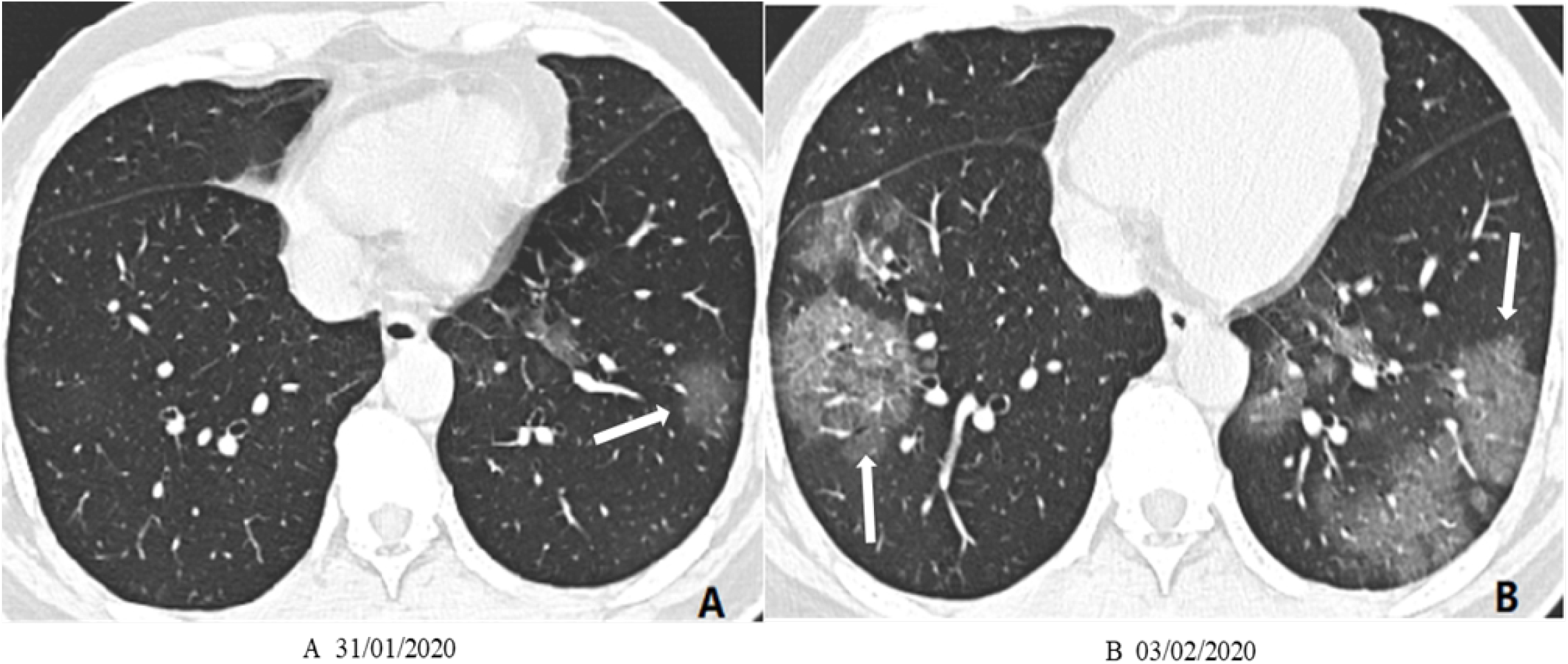
A 44-year-old male patient with COVID-19 pneumonia was admitted to the hospital due to “high fever”. The lung CT at the time of admission (A): GGO of the left lower lobe; high fever persisted after admission and dyspnea occurred. (B) It is suggested that the GGO lesions of both lungs increased significantly and the area increased within 72 hours

**Figure 3:**
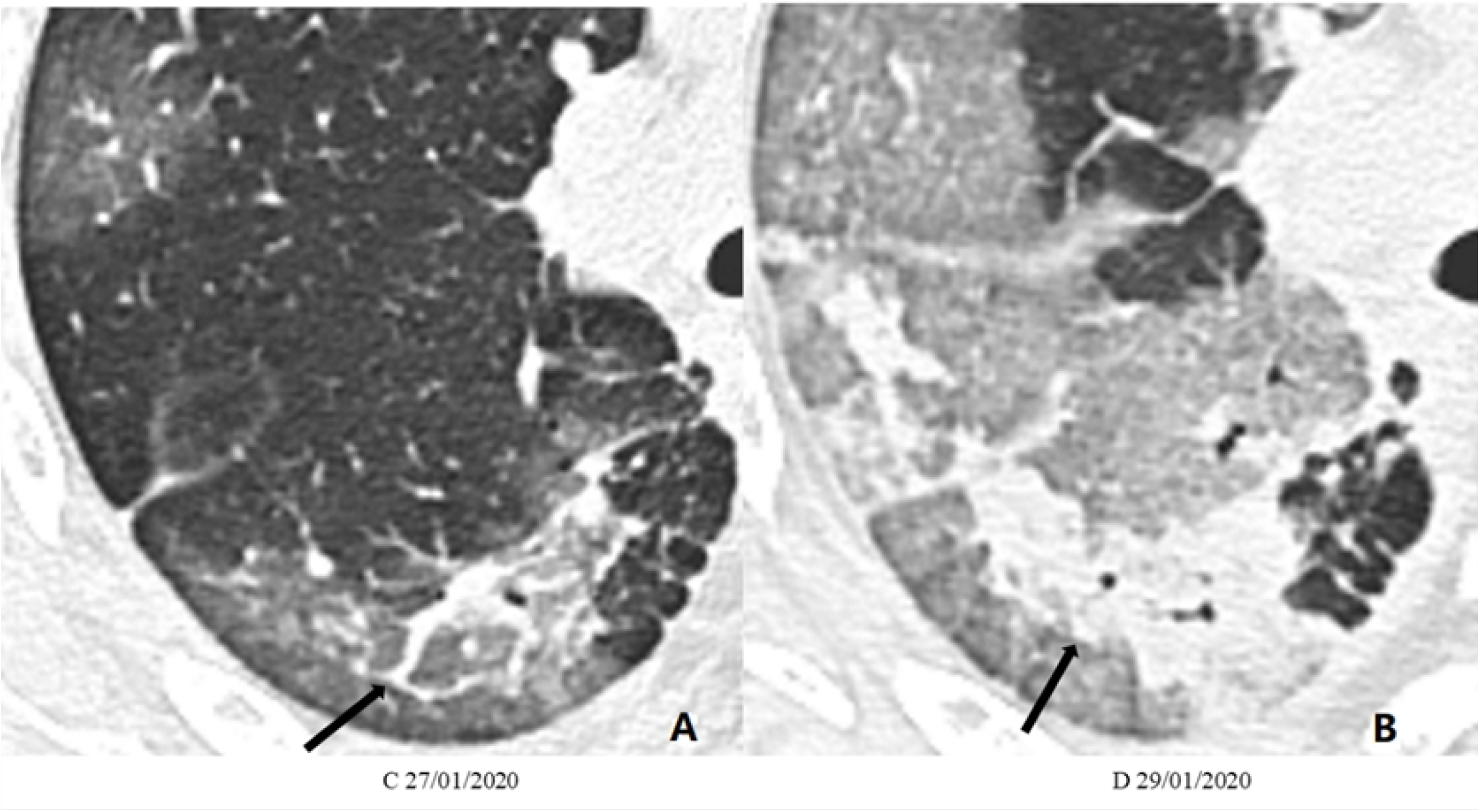
A 60-year-old male patient with COVID-19 pneumonia was admitted to the hospital because of “high fever and dyspnea”. Lung CT (A) showed high-density line-shaped exudation in GGO in the right lower lung. High fever continued after admission With respiratory distress, reexamination CT (B) showed that within 48 hours, the GGO lesions in the lung increased significantly and the area increased, and the high-density linear exudation in the central part of the GGO lesion increased and merged into a sheet.

### 5. Basic principles of early intervention treatment of COVID-19 patients

On the basis of respecting the clinical characteristics and current known pathological process of COVID-19, our team mainly adopts symptomatic supportive treatment measures. Blind medication was avoided during the treatment. All medicines used are considered to effectively protect and support the patient’s own immune function against viruses. As shown in table 5, due to the common hypoproteinemia (89.7%) and hypokalemia (70.1%), we provided corresponding supplements to maintain body fluid electrolytes and internal environment and nutrition balance. Especially in all severe patients, 20-40g albumin / d was given, adjusted according to the heart function. Appropriate plasma supplementation had been applied in 68.8% severe and all critical cases, which could supply immunoglobulins and lymphokines to support the body’s normal antiviral effect. We think that plasma (especially fresh plasma) supplementation has better supportive treatment effects than extracted immunoglobulins. For critically ill patients, an appropriate dose of methylprednisolone had been given to exert anti-inflammatory effects, suppressing the inflammatory storm caused by the body’s excessive immunity. Oxygen therapy had been given in 89.7% of all patients at early stage. When OI declined below 300mmHg, medium-high flow rate to inhale oxygen through the nasal catheter to maintain SaO2 higher than 95%, but if OI declined continually, high-flow nasal cannula oxygen therapy (HFNC) should be applied as early as possible. We found that HFNC could achieve good blood oxygen maintenance effect even in severe and critical patients, with better tolerance than non-invasive mechanical ventilation. With early and appreciated intervention and treatment, only one case took invasive mechanical ventilation for short time.

**Table5.** Early intervention and treatment principles of COVID-19

| Treatment | All patients | Non-severe | Severe(n=26) |  | Analyze |
| --- | --- | --- | --- | --- | --- |
|  |  |  | 150≤OI≤300 | OI≤150 |  |

|  | (n=97) | (n=71) | -mmHg<br>(n=16) | -mmHg<br>(n=10) |  |
| --- | --- | --- | --- | --- | --- |
| Potassium supplement-No., % | 68/97(70.1) | 44/71(62.0) | 14/16(87.5) | 10/10(100) | Maintain normal body fluid environment and electrolyte balance |
| Albumin supplement-No., % | 87/97(89.7) | 61/71(85.9) | 16/16(100) | 10/10(100) | Maintain normal body fluid environment and nutritional status |
| Plasma infusion -No., % | 29/97(29.9) | 8/71(11.3) | 11/16(68.8) | 10/10(100) | Supplement immunoglobulins and lymphokines to support the body's normal antiviral effect |
| Methylprednisolone -No., % | 22/97(22.7) | 6/71(8.5) | 7/16(43.8) | 9/10(90) | Anti-inflammatory, inhibit inflammatory storm, refer to Table 6 for application principles |
| Anticoagulant therapy -No., % | 4/97(4.1) | 0/71 | 1/16(6.2) | 3/10(30) | Prevent thrombosis |
| Antibiotic -No., % | 75/97(77.3) | 49/71(69.0) | 16/16(100) | 10/10(100) | Prevent secondary bacterial infections |
| Nasal catheter oxygen therapy -No., % | 74/97(76.3) | 61/71(85.9) | 13/16(81.3) | 0/10 | Routine and early application in non-critical patients to improve oxygen supply |
| High-flow nasal cannula oxygen therapy (HFNC) | 12/97(12.4) | 0/71 | 3/16(18.8) | 9/10(90) | Oxygenation index nnula oxygen therapy (HFNC)ve oxygen supply 6 for application principlestion index)l "/javascript:;" |
| -No., % |  |  |  |  | rate in d corticosteroid had be recommended not |
| Invasivemechanical ventilation -No., % | 1/97(1.03) | 0/71 | 0/16 | 1/10(10) | After HFNC and methylprednisolone, IO declined continually |

### 6. Application principles and effects of methylprednisolone in severe/critical COVID-19 patients

In this study, 22 patients were treated with methylprednisolone, of which 7 cases were for dealing with high fever in small doses of 40mg. And the other 15 cases were for rescuing, when theyhad sudden respiratory distress and a sharp decrease in OI. In these patients with OI <150mmHg or close to 150mmHg, the application of methylprednisolone varied from 1 to 3 times, but single or as few times as possible application was recommended, rather than continuous and long-term applications. And the single dose of methylprednisolone was 40mg-500mg, according to severity, oxygenation index, speed of progression, production of inflammatory factors, body weight, age and underlying diseases condition. As shown in Figure 4, after treatment with methylprednisolone, the patient’s OI improved significantly, and all but one avoided invasive mechanical ventilation. The most important application principles of methylprednisolone had been shown in table 6.

**Table 6:**
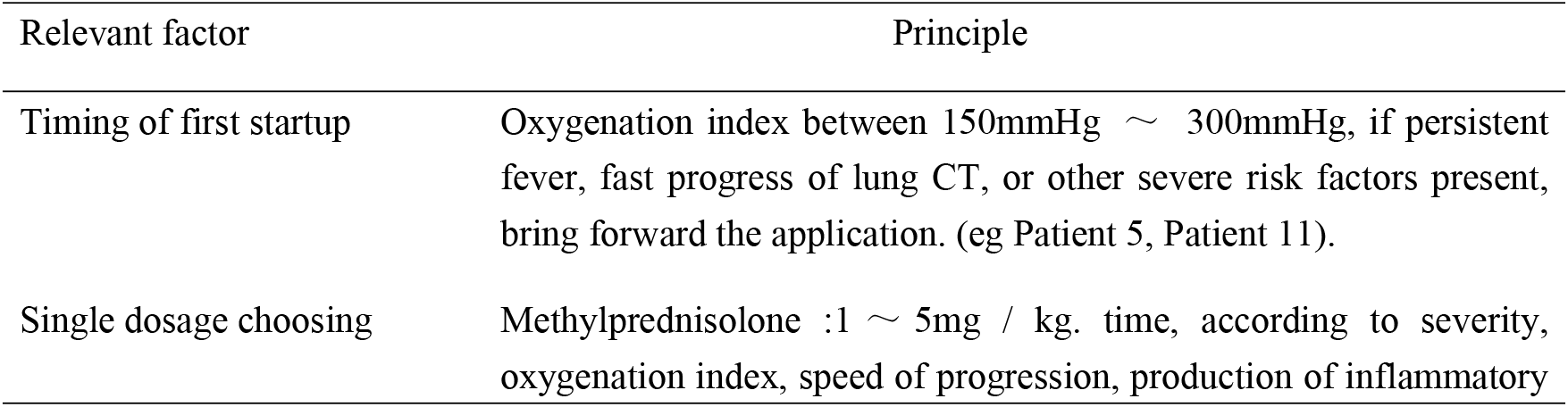
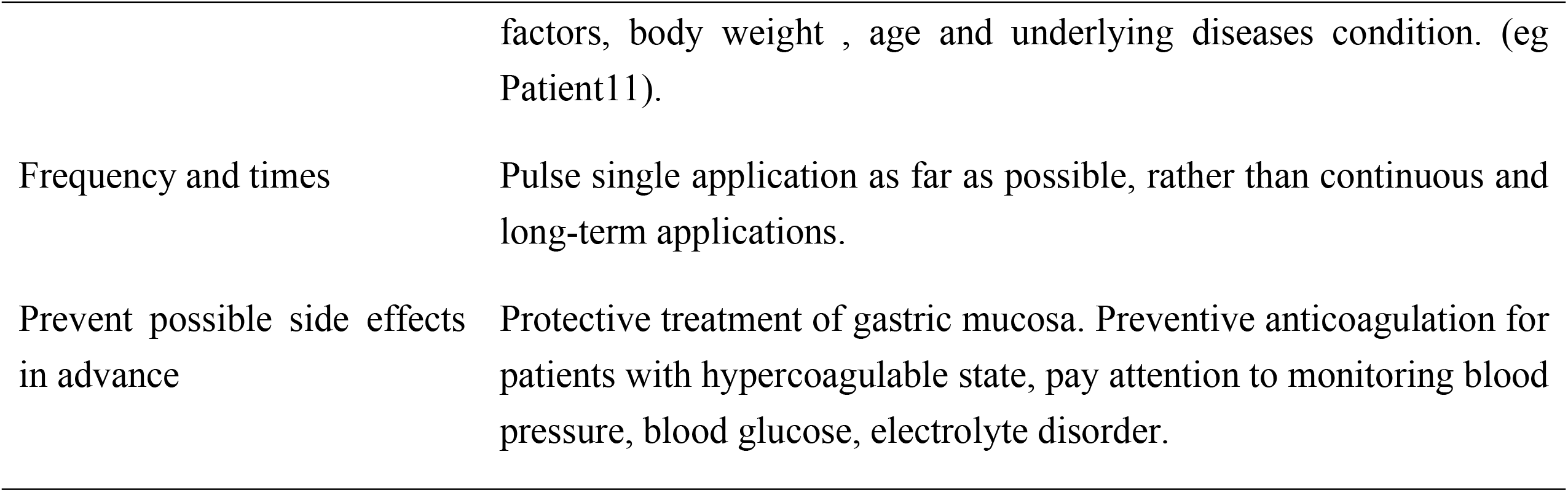
Application principles of methylprednisolone in COVID-19 patients

| Relevant factor | Principle |
| --- | --- |
| Timing of first startup | Oxygenation index between 150mmHg ∼ 300mmHg, if persistent fever, fast progress of lung CT, or other severe risk factors present, bring forward the application. (eg Patient 5, Patient 11). |
| Single dosage choosing | Methylprednisolone :1 ∼ 5mg / kg. time, according to severity, oxygenation index, speed of progression, production of inflammatory |
|  | factors, body weight , age and underlying diseases condition. (eg Patient11). |
| Frequency and times | Pulse single application as far as possible, rather than continuous and long-term applications. |
| Prevent possible side effects in advance | Protective treatment of gastric mucosa. Preventive anticoagulation for patients with hypercoagulable state, pay attention to monitoring blood pressure, blood glucose, electrolyte disorder. |

**Figure 4:**
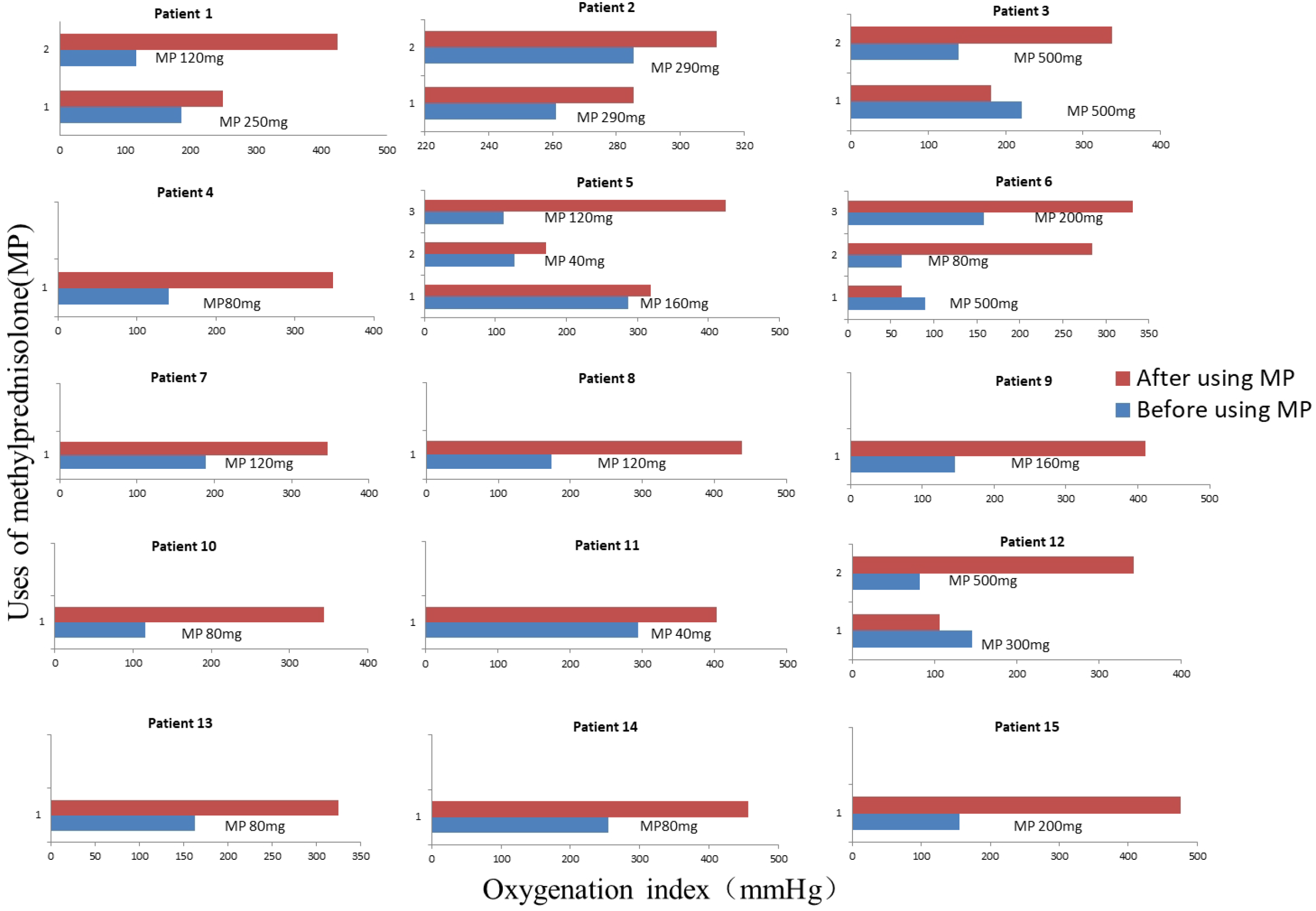
Changes in Oxygenation Index before and after the application of methylprednisolone

Otherwise, it is important to prevent possible side effects in advance, with proton pump inhibitor, appropriated anticoagulation, monitoring blood pressure, blood glucose, electrolyte disorder. All 22 patients treated with methylprednisolone had no gastrointestinal bleeding manifestations; no obvious change in blood glucose and blood pressure, no osteonecrosis of the femoral head (ONFH) in one-month-follow up.

We analyzed the effect of methylprednisolone to time for Negative-conversing in 2019-nCoV testing of nasopharyngeal swab. The time for Negative-conversing in SARS-CoV-2 testing were 10.0 ± 5.3d and 10.0 ± 7.9d in patients with or without methylprednisolone respectively, and there was no statistical difference between the two groups (P> 0.05, Figure 5), indicating that application of methylprednisolone according to above principles does not extend SARS-CoV-2 clearing up time.

**Figure 5:**
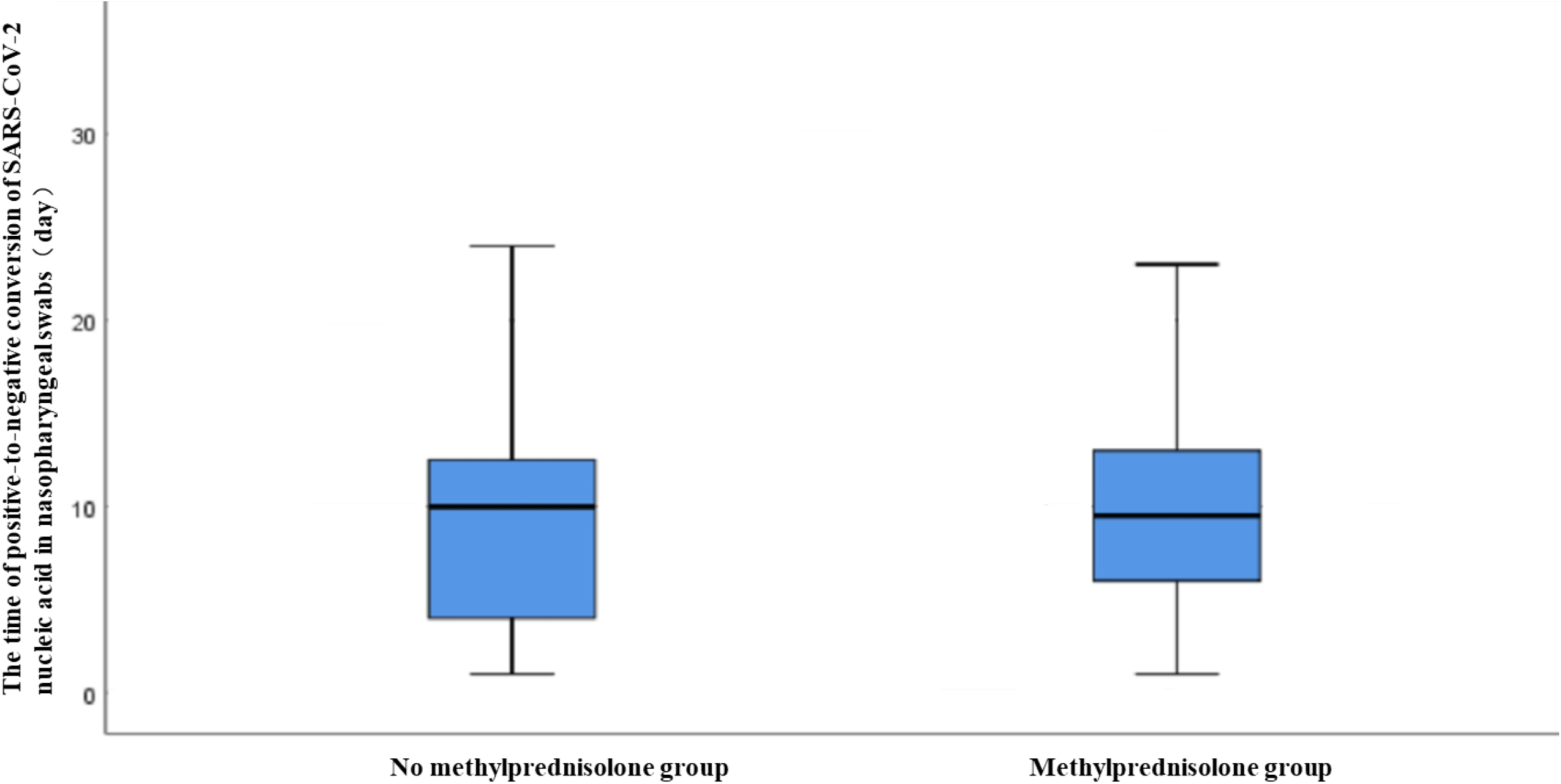
Relationship between methylprednisolone and positive-to-negative conversion of SARS-CoV-2 nucleic acid in nasopharyngeal swabs

## Discussion

COVID-19 is respiratory infectious diseases with global outbreak currently. Up to March 24, 2020, the global case fatality rate was 4.4% ^[2]^. In the cohort study of 41 COVID-19 patients in Wuhan at early stage of epidemic, 13 cases were admitted to the intensive care unit (ICU), and 6 cases died at last ^[3]^. In another cohort study of 138 COVID-19 patients, 36 cases had been transferred to ICU, and of which 4 (11.1%) cases died ^[7]^. The fatality rate was significantly higher in the severe COVID-19 patients. The treatment of mechanical ventilation in critical patients brings also huge medical burden^[4]^. So far, no specific anti-SARS-CoV-2 drug has been found successfully ^[8,9]^. Therefore, it is extremely important to recognize the predictive risk factors of disease aggravation, identify severe disease early, give scientific and effective treatment programs, block the progression to severe disease, and avoid intubation and invasive mechanical ventilation in critical disease to reduce the case fatality rate. Based on successfully treatment of 97 COVID-19 patients (including 26 severe, 10 critical) in our hospital, we would like to share and discuss our experience in recognizing and intervention of severe early and timely.

Consistent with previous research^[10–15]^, persisted or worsen hypoxemia, low lymphocyte count, hypoalbuminemia, hypokalemia, high CRP, high D-dimer, and high BNP were important indicators of predictive risk factors of disease aggravation in our study. Hypoxemia is the most common clinical feature throughout the course of disease (93.8% of all patients) in COVID-19 cases, even if someone have no shortness of breath on admission. The declining of OI in severe patients was more significantly than that in non-severe cases. Shortness of breath was shown in all severe patients, which was 92% of ICU patients in the research of Huang et al^[3]^. A large scale multicenter research ^[12]^ of 1099 COVID-19 patients from 31 provinces reported 95.5%, 81.5% and 59.6% of severe cases had low lymphocyte count, high CRP and high D- dimer respectively, which were significant difference from non-severe ones. In the research of Huang^[3]^, low lymphocyte count, hypoalbuminemia and high D- dimer had been found common in severe COVID-19 patients, particularly in critical cases in ICU. In addition, we also found the hypokalemia and high BNP were more significant in severe cases than non-severe ones. In addition, SARS-CoV-2 viremia should also be paid attention to, as a predictor for progression to severe. Except for the abnormal values of above indicators, we found that it was more important to observe the changing trend of these indicators during course of disease. More significant low lymphocyte count and high CRP appeared in severe patients from the beginning and continued the whole course, which indicated the imbalance between early immune response and lateral inflammatory reaction. Liu^[11]^ et al reported that mean days from illness onset to ICU admission were 8d in the severe or critical type. In our study, the peak of obvious abnormality of D-dimer and BNP appeared at day 8-9 of the disease course and then decreased slowly, which indicated early recognition of critical signs and the risk of secondary thrombosis and myocardial injury. The persistent hypoalbuminemia and hypokalemia suggested that the imbalance of homeostasis, at the same time instructed treatment.

Some characteristic imaging findings, such as GGO, nodules, stripe and patchy consolidation, bronchial abnormalities, were consistent with previous studies ^[3,12,16]^. The abnormal images on lung CT were corresponding to the pathological features of COVID-19 pneumonia^[17]^. Two types of acute exudative pathological changes had been considered corresponding to imaging change in these COVID-19 patients: mainly serous exudation (mainly albumin exudation and manifested as GGO in lung CT), and fibrinous exudation (fibrin exudation and manifested as stripe and patchy consolidation in lung CT). Otherwise, bleeding exudation might happen on a few severe patients with hemoptysis, but no obvious damage of lung structure showed in lung CT. As for the characteristic imaging findings and the progress of imaging change on lung CT in COVID-19 patients, it is more meaningful to judge the severe risk of the disease. In my opinion, lung CT should be reviewed and evaluated timely. Multi-lobar or rapidly progressing GGO and consolidation on chest CT were important indicators of predictive risk factors of disease aggravation in our study.

According to the clinical characteristics of COVID-19, early and timely intervention is important. In this study, none of the 97 patients had basic diseases of immune dysfunction. Therefore, it is considered that immune dysfunction and internal environmental disorder are transient after the virus infection. So in the treatment, we mainly focused on symptomatic and supportive treatment in the early stage, such as adequate supplementation of albumin, plasma and potassium, in order to maintain the stability of the intracellular environment and adequately reactivate body immunity to clean up sars-cov-2 by themselves, which is consistent with Lei Zhang’s opinion ^[18]^. At the same time, continuous application of small doses of glucocorticoid should be avoided in case of suppressing the body’s immune function at this stage.

HFNC should be applied as early as possible, if patients had progressive decline of OI. We found that HFNC could achieve good blood oxygen maintenance effect even in severe and critical patients, with better tolerance than non-invasive mechanical ventilation. The most common indications for invasive mechanical ventilation should be strictly control in critical COVID-19 patients. Based on our experience, appropriate pulse methylprednisolone application on severe/critical COVID-19 patients on demand, improved blood oxygen and reduced the utilization rate of invasive mechanical ventilation, case fatality rate and medical burden significantly. A meta-analysis ^[19]^ shows that glucocorticoid is a more effective drug for suppressing the occurrence of inflammatory storms, and it is safe within a certain dose range. The emergence of COVID-19 inflammatory storm is often life-threatening in a short time, with a variety of inflammatory factors^[3]^, so the effect of single IL-6 mAb on COVID-19 should be further evaluated ^[20]^. If the inflammatory storms could not be suppressed timely, multiple organ failure and death might be caused. So far, glucocorticoid may be unique effective medicine suppressing the progress of the inflammatory storm with appropriated and reasonable application. The application of glucocorticoids in viral pneumonia is currently controversial. A retrospective cohort study about SARS showed that glucocorticoids can reduce case fatality rate and hospital stays ^[21]^, but there are also studies that show that glucocorticoids may increase the case fatality rate of SARS patients and delay the virus clearance time ^[22, 23]^. In patients with Middle East Respiratory Syndrome (MERS) and influenza virus pneumonia, the use of glucocorticoids is also controversial ^[24,25,26,27,28]^. Application indication and dosage of corticosteroid had been strictly controlled from the beginning of epidemic of COVID-19, and corticosteroid had be recommended not be used for the treatment of 2019-nCoV-induced lung injury or shock outside of a clinical trial ^[29]^. Although this issue has always been controversial ^[30]^, clinical usage of corticosteroid was obviously restricted, and the role was underestimated greatly.

The most obvious effects of methylprednisolone therapy in our study were that invasive mechanical ventilation had been avoided in 14 patients, the time to use invasive mechanical ventilation had been shortened significantly in one patient, medical expenses had been reduced greatly, and no death happened. There were no obvious adverse reactions during one-month follow up. According to some reports, we find that there are too many critical patients admitted in ICU at the same time or there is no qualification or condition of invasive mechanical ventilation treatment in some hospital admitting COVID-19 patients. In this case, methylprednisolone therapy might be an effective choice. Based on the data and experience of our team, it is recommended to improve the status of glucocorticoids in the treatment of severe COVID-19, and organize multi-center large-scale retrospective studies and possible prospective studies.

## Conclusion

After summarizing the successful treatment of 97 COVID-19 patients (26 severe cases) by our team, we found accurate and timely identification of clinical features in severe risks, and early and appropriate intervention could block disease progression. Appropriate dose of methylprednisolone can effectively avoid invasive mechanical ventilation and reduce case fatality rate in severe and critical COVID-19 patients.

## Data Availability

The readers can access the data supporting the conclusions of the study in the manuscript and updated manuscript if the data has been updated

## Acknowledgments

The authors would like to thank the study participants who donated their samples for the advancement of scientific knowledge.

## Author contributions

Experimental conception and design: Jin Huang, Jing Liu, Hong Shan, Meizhu chen, Changli Tu. Collection of samples: Meizhu chen, Changli Tu, Cuiyan Tan, Xiaobin Zheng, Xiaohua Wang, Jian Wu, Yiying Huang, Zhenguo Wang, Yan Yan, Zhonghe Li. Performing the experiment: Meizhu Chen, Changli Tu, Cuiyan Tan. Data analysis: Changli Tu, Meizhu Chen. Contribution of reagents, materials and/or analysis tools: Changli Tu, Meizhu chen, Jin Huang, Jing Liu. Writing the paper: Meizhu Chen, Changli Tu. Critical review and approval: Jin Huang, Jing Liu, Hong Shan.

